# Strengthening chlamydia management in Australian general practice using interventions that align with the clinic workflow: implementation and feasibility trial findings

**DOI:** 10.1101/2025.11.17.25340442

**Authors:** Jane L Goller, Zoie Alexiou, Helen Bittleston, Meredith Temple-Smith, Lena Sanci, Julie Simpson, Marcus Y Chen, Christopher K Fairley, Deborah Bateson, Christopher Peter Bourne, Natalie Carvalho, Basil Donovan, Stephanie C Munari, Jane Tomnay, Rebecca Guy, David Regan, David Hawkes, Marion Saville, Claudia S Estcourt, Jane S Hocking, Jacqueline Coombe

## Abstract

**Objective:** To evaluate the implementation and impact of the Management of Chlamydia Cases in Australia (MoCCA) intervention for strengthening chlamydia management in Australian general practice.

**Design:** Non-randomised implementation and feasibility trial

**Setting:** General practices in the Australian states of New South Wales, Queensland and Victoria.

**Participants:** 14 general practices participated and implemented the MoCCA intervention.

General practitioners (GPs) and nurses from participating clinics were interviewed about implementation and use of MoCCA. Deidentified patient attendance and clinical data for patients aged 16-44 years were collected for each clinic.

**Intervention:** Multifaceted intervention (website (www.mocca.org.au/), patient factsheets, shortcuts for documenting a chlamydia consultation, mailed specimen kits for chlamydia retesting, guidance articles, continuing professional development activities) to facilitate chlamydia management in general practice. Clinicians (GPs and nurses) were asked to use the MoCCA intervention to guide their chlamydia and pelvic inflammatory disease (PID) diagnosis and management over a 12-month intervention period.

**Main outcome measures:** The primary outcomes focussed on intervention implementation (acceptability, adoption, appropriateness, feasibility, fidelity, penetration, sustainability). Secondary outcomes focused on chlamydia retesting and reinfection among patients who had a chlamydia infection and PID rates among female patients, comparing the intervention period with a 12-month preintervention period.

**Results:** Clinicians (doctors=26, nurses=7) largely viewed MoCCA as compatible (**appropriateness**) with the general practice setting and an **acceptable** approach to streamlining chlamydia management. Some MoCCA components (e.g. website, shortcuts, factsheets, patient delivered partner therapy [PDPT] resources) were preferred for their ease of use and support for decision making over other components (e.g. postal retesting kits). Intention to use MoCCA (**adoption)** and its uptake (**penetration**) varied between clinicians and MoCCA components. Some clinicians intended to use MoCCA but did not due to lack of opportunity, forgetting, or other preferred practices. MoCCA was not used uniformly (**fidelity**), but its flexible design let clinicians use MoCCA components to suit their needs. Clinicians liked that MoCCA facilitated best practice chlamydia management (**feasibility**) as outlined in Australian STI management guidelines. Reported improvements in care included better quality, continuity and time-efficiency (e.g. website simplified chlamydia management) and patient communication (e.g. shortcuts/factsheets remind to discuss retesting). Many wanted access to MoCCA resources post-study (**sustainability)**. Chlamydia tests were conducted in 17.2% patients preintervention (n=7670/44607, with 4.6% positive) and 18.0% (n=9703/54008, with 5.0% positive) during the intervention. Proportions retested were 18.1% (95% confidence interval (CI) 13.4, 23.7) preintervention and 23.1% (95%CI 18.7, 28.1) in the intervention period (difference=5.0%, 95%CI −2.1, 12.1). Adjusting for gender and clinic location (metropolitan vs non-metropolitan), retesting for 16–25-year-olds increased between the preintervention and intervention period (adjusted relative risk (aRR) 2.12, 95%CI 1.43, 3.15) and for individuals aged 26-44 years results were inconclusive (aRR 1.01, 95%CI 0.66,1.55). PID diagnosis per 1000 consultations among females were 16.9 preintervention and 19.1 intervention period (absolute difference = 2.18%, 95% CI 0.13, 4.23).

**Conclusions:** MoCCA aligned with the general practice setting and supported improved chlamydia management.

## Background

*Chlamydia trachomatis*, the pathogen responsible for chlamydia, is the most prevalent bacterial sexually transmitted infection (STI) globally and in Australia.^1, 2^ Although usually asymptomatic, chlamydia can cause serious long-term complications particularly among individuals with female reproductive organs. These include pelvic inflammatory disease (PID), ectopic pregnancy, and infertility.^3^ Repeat infections increase the risk of PID, particularly in younger individuals.^4^

Historically, chlamydia control efforts around the world have focused largely on screening of asymptomatic individuals.^5^ However, there is increasing recognition of the limitations of widespread testing to reduce chlamydia prevalence and its complications.^6-8^ Chlamydia control efforts in many countries including the UK, Australia and the Netherlands now increasingly focus on management of diagnosed infections to reduce the risk of reproductive sequelae.^5, 9^ For example, in Australia, the National STI Strategy has reduced its focus on test uptake and places increased emphasis on strengthening management of diagnosed infections, particularly on reducing repeat infections and early PID detection.^10^ General practice guidelines now recommend opportunistic chlamydia testing only for sexually active women aged under 25 years or for women >25 years if at increased risk.^11^

General practice is Australia’s mainstream primary care setting and where most chlamydia infections are diagnosed and treated.^12, 13^ As such, it is a crucial setting for optimising chlamydia management. However, STIs are one of a myriad of health conditions that general practitioners (GPs) encounter in their routine clinical practice^14^ and there are barriers to optimal care including lack of sexual health expertise and time among GPs and patient factors such as confidentiality concerns.^15-17^ Nurses are a vital part of the Australian general practice workforce, contributing to many healthcare areas including immunisation, chronic disease management, lifestyle education ^18^ and in some practices, sexual health care.^19^ Most diagnosed chlamydia infections in Australia are treated with antibiotics ^20, 21^ but high reinfection rates (up to 22%)^22^ suggest an index case’s sexual contacts are not always informed or treated. Australian STI Management Guidelines^23^ recommend partner management for sexual contacts of those diagnosed with chlamydia and retesting after treatment to check for chlamydia reinfection at 3 months after treatment, but retesting rates remain low (<25% where measured).^21, 24^ Further, in Australian general practice, low rates of PID diagnosis have been reported suggesting it is underdiagnosed in this setting^6^ and there is evidence that GPs encounter barriers to conducting pelvic examinations for women experiencing pelvic pain, potentially impacting capacity to diagnose PID.^25^

The Management of Chlamydia Cases in Australia (MoCCA) study aimed to develop a scalable intervention for strengthening chlamydia management in Australian general practice, with a particular focus on i) improving partner management, including use of patient delivered partner therapy (PDPT) where relevant; ii) timely retesting for chlamydia reinfection and; iii) early and improved detection of PID.^26^ PDPT is where the diagnosing clinician provides a prescription or course of antibiotic treatment to the index case for treatment of their partner/s, without seeing the partner, thereby expediting partner treatment.^27^ MoCCA was a partnership project, involving researchers and 11 partner organisations (e.g. health departments, clinical services) with a shared vision to strengthen chlamydia management in general practice. This partnership provided a structure that facilitated clinical and strategic contributions to developing the MoCCA intervention and ensuring the study aligned with Australian priorities for STI control, chlamydia management and the general practice setting. In the early stage of MoCCA, we conducted surveys, interviews and focus groups with clinicians, health consumers and policy makers to understand the context and needs for chlamydia management in general practice.^25, 28-31^ We found a need for more information and resources for clinicians and patients about best practice chlamydia management and, crucially that our intervention must integrate into the general practice setting. Subsequently, we developed a multi-faceted intervention to facilitate chlamydia management that we piloted and then refined.^26^

^32^ Our refined MoCCA intervention (Box 1) comprises a website (www.mocca.org.au), patient factsheets, shortcuts for documenting a chlamydia consultation in the electronic medical record (EMR)^32^, mailed specimen kits for chlamydia re-testing, guidance articles^27, 33^ and continuing professional development (CPD) activities. During 2023-2024, we evaluated the MoCCA intervention in Australian general practice in a non-randomised implementation and feasibility trial.^26^ In this mixed methods study we report on the implementation and impact outcomes of this trial.

### Box 1: Overview of the MoCCA intervention

- ***Website***(www.mocca.org.au) is the central resource that outlines best practice chlamydia management and links to key Australian guidelines and resources and provides:
  - Clinical decision-making resources, flowcharts and tips for discussing partner notification, retesting and for PDPT
  - Published education articles and CPD activities
  - Information about PID and epididymo-orchitis
  - Brief information about the diagnosis and management of other STIs (gonorrhoea, syphilis, *Mycoplasma genitalium)*.
- ***Patient factsheets***with information for patients about chlamydia, notifying partners, retesting and PID.
- ***Shortcuts***for streamlining documentation of a consultation involving: i. chlamydia management, ii.
- PID management, and iii. provision of PDPT. These are imported into the EMR and provide editable text populated in patient clinical notes by typing the shortcut text.
- ***Flowcharts***detailing information about PDPT eligibility, PID diagnosis and options for organising retesting.
- ***PDPT prescription template***for import into the EMR.
- ***Mailed specimen kits***for chlamydia retesting to check for reinfection.
- ***Evidence-based guidance articles***that outline best practice support for chlamydia management, PID management and for PDPT decision making.
- ***CPD activities***that aligned with the MoCCA study.

## Methods

We conducted a non-randomised implementation and feasibility trial to evaluate the MoCCA intervention in Australian general practice. Our detailed trial methods are described in the published protocol.^26^ We implemented MoCCA in 14 general practices located in the Australian states of New South Wales (NSW), Queensland (QLD) and Victoria (VIC). Participating practices (clinics) were recruited via advertisements in general practice networks or directly via phone and email.

Researchers directly supported clinics to set up the MoCCA intervention before the trial commenced. This included: bookmarking the MoCCA website on the clinics’ internet browser, importing documentation shortcuts and a PDPT prescription template into EMR software, setting up postal retesting, and making patient factsheets accessible to clinic staff, with each component taking from <5 to at most 30 minutes to set up. Resources (desktop notices, quarterly emailed newsletters) to support adoption and engagement were provided to remind clinicians of MoCCA resources (e.g. website address). Clinicians (GPs and nurses) were asked to use MoCCA to guide their chlamydia and PID diagnosis and management over a 12-month intervention period. A formal induction meeting was held with each clinic during which the intervention was demonstrated and strategies to embed it into routine clinical practice were discussed.

### Outcomes and data collection

Our primary trial outcomes (Box 2) focused on success of implementing MoCCA^26^ and included acceptability, adoption, appropriateness, feasibility, fidelity, penetration, and sustainability.^34^ We conducted semi-structured interviews with clinic staff at the three month point of the intervention to understand barriers and facilitators to implementation (results published separately).^35^Toward conclusion of the intervention period (9–12-month point) and to assess our implementation outcomes we conducted further semi-structured interviews and a workshop with clinical staff from participating clinics. Interviews and the workshop were conducted by HB, JG, JC and MTS using a semi-structured guide developed by MoCCA researchers (supplemental information). Questions focused on the participants’ knowledge and use of MoCCA, what helped or hindered its use, the impact of MoCCA (if any) on clinical practice and how/if MoCCA should be modified for use post-trial. A wrap-up meeting was also held with each clinic to communicate how to finish the study and as further opportunity for clinic staff to provide feedback about MoCCA. Detailed meeting notes were taken. Although cost is a further implementation outcome,^34^ we do not report implementation costs due to our focus on the concluding stages of the intervention and on understanding how MoCCA was or was not used to support chlamydia management.

#### Box 2: Definition of MoCCA implementation outcomes

1. **Acceptability** – extent that the content of the MoCCA intervention and complexity of its implementation and use is agreeable to the general practice setting.
2. **Adoption** – readiness to use and uptake of the MoCCA intervention.
3. **Appropriateness** - relevance of the MoCCA intervention to the general practice setting and to support chlamydia management.
4. **Feasibility** - extent that the MoCCA intervention can be successfully used in general practice to support chlamydia management.
5. **Fidelity** – extent that the MoCCA intervention was implemented and used as intended.
6. **Penetration** – extent the MoCCA intervention is used during the trial and throughout the clinic.
7. **Sustainability** – extent that intended use of the MoCCA intervention was maintained. Opportunities for real-world access to the MoCCA intervention.

Invitations to participate in an interview or workshop were sent by email to the key contact person from each clinic and included in emailed study newsletters. Those who expressed interest were emailed a plain language statement about the study and a mutually suitable time was arranged. The interviews and workshop were conducted, audio-recorded, and transcribed verbatim using videoconferencing and transcription software with verbal consent obtained before commencing the recording. Transcripts were verified, cleaned and deidentified by HB and JG. Transcripts were stored in a password-protected secure server at the University of Melbourne. Interview participants were provided a $100 voucher and workshop participants a $150 voucher as reimbursement for their time.

We also explored the impact of MoCCA on chlamydia management outcomes. Our primary impact outcome was retesting within 2 to 4 months (to detect retesting at the recommended 3 month interval) of a chlamydia infection (defined as date of positive test).^23^ Other impact outcomes were PID rates among female patients, and, reinfection rates among patients who had a chlamydia infection. As a post-hoc additional secondary impact outcome, we examined retesting within 2months, in consideration that retesting too early can detect remnant chlamydial nucleic acid, giving a false positive diagnosis.^36^

Deidentified patient attendance and clinical data were collected from the clinic EMR using GRHANITE (www.grhanite.com/) data extraction software and used to measure retesting, reinfection and PID rates. Data-items included visit date, clinic, test request, test result, age at visit, gender, and clinical encounter reason. These data were collected for the 12-month pre-intervention period (defined as 12 months before a clinic’s intervention start date) and the intervention period (up to 31st July 2024) for patients aged 16-44 years who had a consultation at any point during this period. These periods were individualised for each clinic based on its intervention start and end dates.

### Patient involvement

As stated above, MoCCA was a partnership between researchers and a group of organisations including clinical services and health departments (representing patients, public and healthcare providers) that contributed to the design and conduct of all stages of the project.

## Data analysis

Assessment of our implementation outcomes was guided by two complementary frameworks, the Consolidated Framework for Implementation Research (CFIR)^37^ and Normalisation Process Theory (NPT)^38^ that can support understanding of the complexities of integrating interventions into routine practice.^39^ The CFIR encompasses five domains (intervention characteristics, outer setting, inner setting, characteristics of individuals, and processes) of which each domain contains several constructs. It can be used to understand the contextual elements that influence implementation.^37^ NPT comprises four constructs (coherence, cognitive participation, collective action and reflexive monitoring) that can help in understanding the cognitive and social processes used by individuals to integrate an intervention into practice.^38^

Qualitative analysis was conducted in NVivo 14 (Lumivero 2023, www.lumivero.com) and commenced with data familiarisation via reading transcripts, noting preliminary observations and discussion with authors (JG, HB, JC). We then established a coding framework across the five CFIR domains (and its sub-constructs) and the four NPT constructs and undertook a directed content analysis of transcript data in relation to these frameworks. We present findings for our implementation outcomes^34^ and identify how the CFIR and NPT helped to understand these. Interview and workshop transcripts were analysed firstly followed by review of our clinic meeting notes to confirm and clarify our findings.

Quantitative patient attendance and clinical data were managed in StataSE 18 (Stata, College Station, TX, USA) and analysed in R (V4.3.0). For our impact outcomes, retesting was calculated as the proportion of individuals with chlamydia who had a retest conducted or requested within i) 2– 4 months and ii) 7 days to <2 months after their initial diagnosis. We limited data for our retesting and reinfection analysis to patients who had a first positive test in the first 8 months of the pre-intervention or intervention period to allow time for follow up. Reinfection rates were determined as the proportion of individuals who, upon retesting, tested positive for chlamydia. PID rates were calculated as the proportion of women aged 16–44 years with consultations that resulted in a PID diagnosis. We defined PID as ‘definite’ based on reason for clinical encounter of PID or salpingitis in the EMR or as ‘probable’ based on clinical encounter reason of cervicitis, endometritis, dyspareunia, pelvic pain, or post-coital bleeding as well as abdominal pain in the presence of an STI or STI test including for chlamydia, gonorrhoea, *Mycoplasma genitalium*. We also calculated chlamydia testing rates between our 12-month pre-intervention and intervention period as the proportion of clients attending in each period who had one or more chlamydia test requested. Positivity was the proportion of individuals with a chlamydia test (including retests) that had a positive result in any of the tests performed (e.g. if a patient was tested from multiple anatomical sites on the same day, all tests and positive results were collapsed and counted once).

We compared our retesting and PID outcomes between our 12-month pre-intervention and intervention period, absolute differences and 95% CIs were calculated. Poisson regression models, with a binary indicator for intervention and adjustment for potential confounders were used to estimate the relative impact of the overall MoCCA intervention (presented as a relative risk (95% CI)). Robust standard errors were used to account for clustering by clinic. A priori, we defined potential confounders as patient gender, age at consult and clinic location (metropolitan versus non-metropolitan).^4 24 40^ The intracluster correlation coefficient (ICC) was estimated from a separate mixed-effects Poisson model with no covariates and used to quantify within-clinic similarity in outcomes. We tested whether intervention effects differed by patient gender, age at consult and clinic location using interaction terms.

## Results

The MoCCA implementation and feasibility trial spanned from January 2023 to August 2024 in 14 clinics located in NSW (n=5), QLD (n=2) and VIC (n=7) (Table 1). Based on postcode, 10 clinics were located in metropolitan areas and four in more disadvantaged areas of Australia. The largest clinic employed 50 staff including 15 GPs and 8 nurses. The smallest clinic employed six staff, including three GPs and no nurses. Five clinics had a strong focus on sexual health care, one largely cared for homeless people and another largely for international students.

**Table 1:** Characteristics of participating general practice clinics, n=14.

| Total clinics (N=14) |  | n | % |
| --- | --- | --- | --- |
| State / Territory | New South Wales | 5 | 35.7 |
|  | Queensland | 2 | 14.3 |
|  | Victoria | 7 | 50.0 |
| Clinic location* | Metropolitan | 10 | 71.4 |
|  | Non-metropolitan | 4 | 28.6 |
| Socio-economic advantage and disadvantage, quintile# | 5 (most advantaged) | 6 | 42.9 |
|  | 4 | 3 | 21.4 |
|  | 3 | 1 | 7.1 |
|  | 2 | 2 | 14.3 |
|  | 1 (most disadvantaged) | 2 | 14.3 |
| **Billing<sup>^</sup>** | Private | 2 | 14.3 |
|  | Mixed | 9 | 64.3 |
|  | Bulk billing | 3 | 21.4 |
| **Number of GPs** | 1-5 | 4 | 28.6 |
|  | 6-10 | 7 | 50.0 |
|  | 11+ | 3 | 21.4 |
| **Number of nurses** | 0-2 | 7 | 50.0 |
|  | 3-5 | 6 | 42.9 |
|  | 6+ | 1 | 7.1 |

\* Using Modified Monash Model classification: (https://www.health.gov.au/resources/apps-and-tools/health-workforce-locator/app), non-metropolitan includes regional towns, large and small rural towns. #Using the Index of Relative Socio-economic Advantage and Disadvantage for the postcode of the clinic. <sup>^</sup>Bulk billing refers to when the healthcare provider bills Medicare (Australia's universal insurance scheme) directly for the clinical consultation, and the patient does not incur any out-of-pocket costs.

### Attendance and testing

Within these clinics, the MoCCA intervention had a mean duration of 12.8 months (SD 1.6, range 8.6-14.8), during which 54,008 individuals aged 16-44 years attended for a total of 263,114 visits. The pre-intervention period was 12 months during which 44,607 individuals attended for a total of 200,142 visits. Among these patients, a total of 17,373 chlamydia tests were conducted representing 17.5% of all patients who tested once or more across both periods [pre-intervention n=7670 (17.2%), intervention n=9703 (18.0%)]. The characteristics of patients tested and the proportion positive by intervention period is provided in table 2. Among patients tested, 4.6% (n=353) were positive for chlamydia in the pre-intervention period and 5.0% (n=482) in the intervention period. Positivity was higher for males (9.2% pre-intervention, 8.4% intervention) than females (3.7% pre-intervention, 4.2% intervention). Data for patients with a positive test were included in our retesting and reinfection analyses and data for all women attending formed our dataset for assessing PID rates.

**Table 2:** Chlamydia testing and positivity by intervention period.

|  | Pre-intervention# |  | Intervention* |  |
| --- | --- | --- | --- | --- |
|  | Tests/Patients<br>n/N (%) | Positive/Patients tested<br>n/N (%) | Tests/Patients<br>n/N (%) | Positive/Patients tested<br>n/N (%) |
| **Total** | 7670/44607 (17.2) | 353/7670 (4.6) | 9703/54008 (18.0) | 482/9703 (5.0) |
| **Clinic location** |  |  |  |  |
| Metropolitan | 6476/34761 (18.6) | 286/6476 (4.4) | 8319/43340 (19.2) | 433/8319 (5.2) |
| Non-metropolitan | 1194/9846 (12.1) | 67/1194 (5.6) | 1384/10668 (13) | 49/1384 (3.5) |
| **GPs at clinic** |  |  |  |  |
| 1-5 | 4107/20676 (19.9) | 142/4107 (3.5) | 5149/25799 (20.0) | 205/5149 (4.0) |
| 6-10 | 2370/14690 (16.1) | 178/2370 (7.5) | 1540/10234 (15.0) | 228/3014 (7.6) |
| 11+ | 1193/9241 (12.9) | 33/1193 (2.8) | 3014/17975 (16.8) | 49/1540 (3.2) |
| **Gender** |  |  |  |  |
| Female | 6400/32063 (20.0) | 237/6400 (3.7) | 7960/40145 (19.8) | 334/7960 (4.2) |
| Male | 1252/12341 (10.1) | 115/1252 (9.2) | 1716/13613 (12.6) | 145/1716 (8.4) |
| Other | 18/203 (8.9) | 1/18 (5.6) | 27/250 (10.8) | 3/27 (11.1) |
| **Age group** |  |  |  |  |
| 16-19 years | 537/3176 (16.9) | 42/537 (7.8) | 559/3472 (16.1) | 50/559 (8.9) |
| 20-24 years | 2025/8827 (22.9) | 115/2025 (5.7) | 2457/10637 (23.1) | 159/2457 (6.5) |
| 25-29 years | 2240/9870 (22.7) | 85/2240 (3.8) | 2852/12639 (22.6) | 122/2852 (4.3) |
| 30-44 years | 2868/22734 (12.6) | 111/2868 (3.9) | 3835/27260 (14.1) | 151/3835 (3.9) |

#Pre-intervention is defined as the 12 months prior to the start of the intervention. \*Intervention is defined as the full intervention period.

**Table 3.**
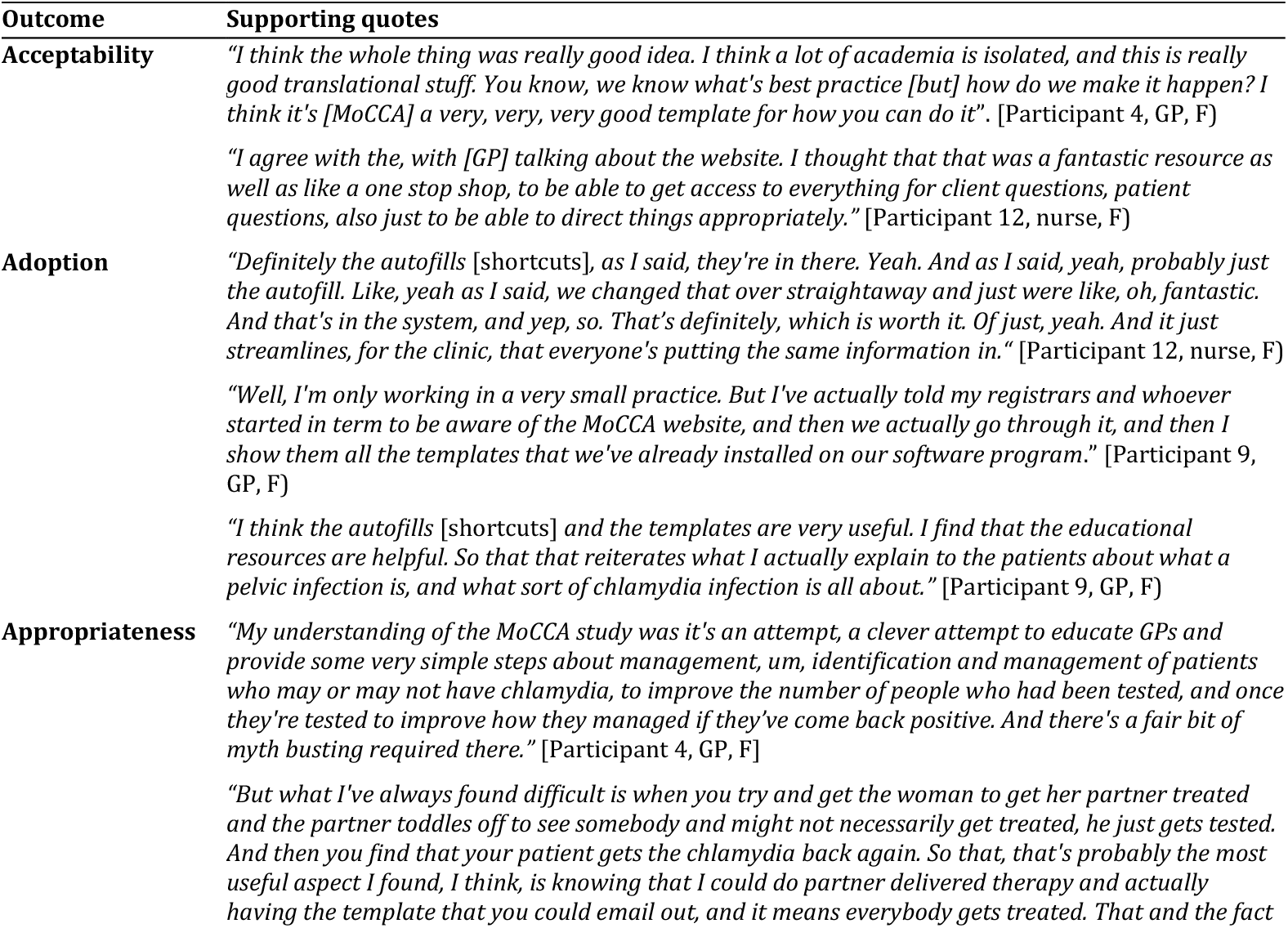
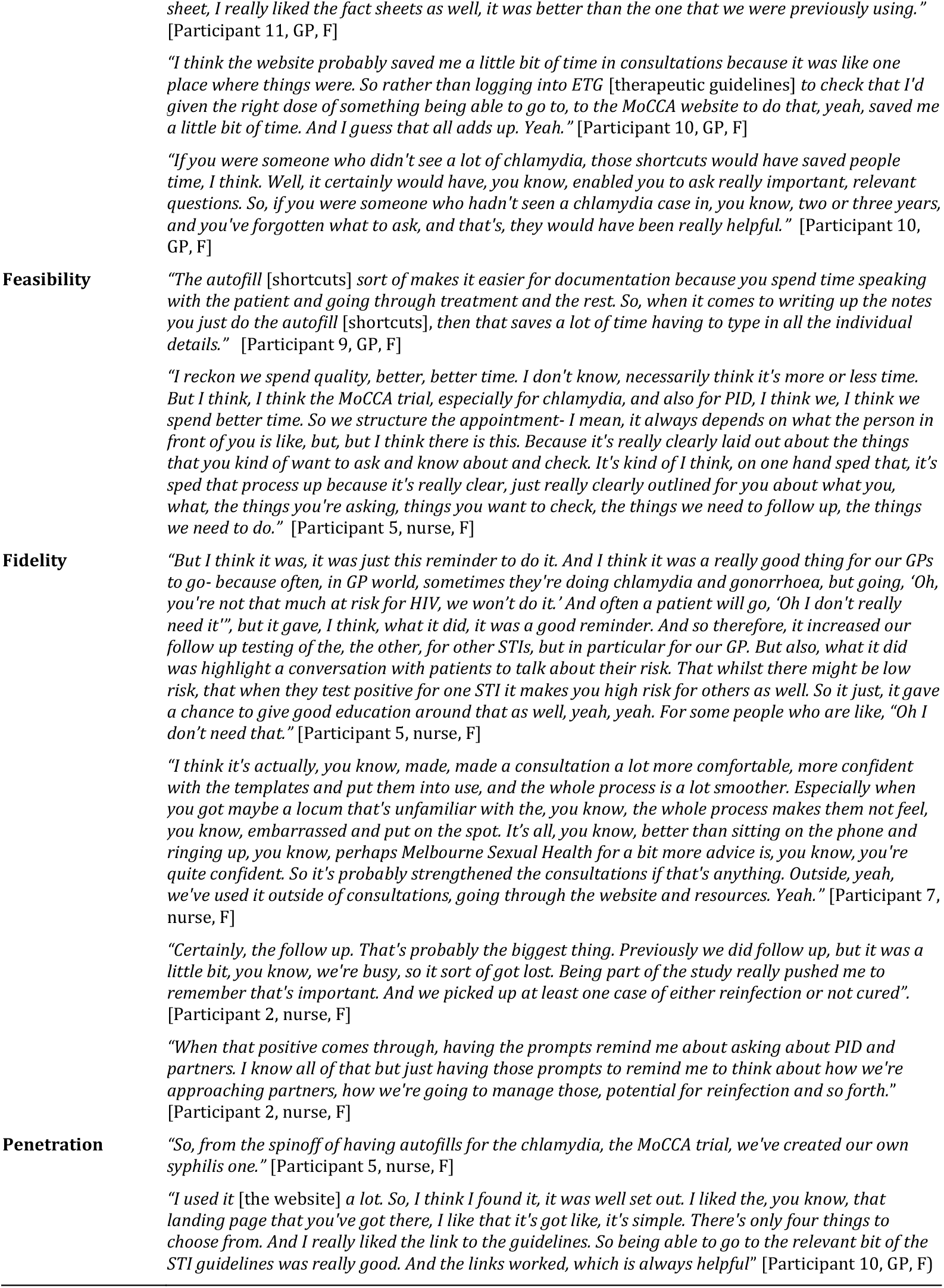
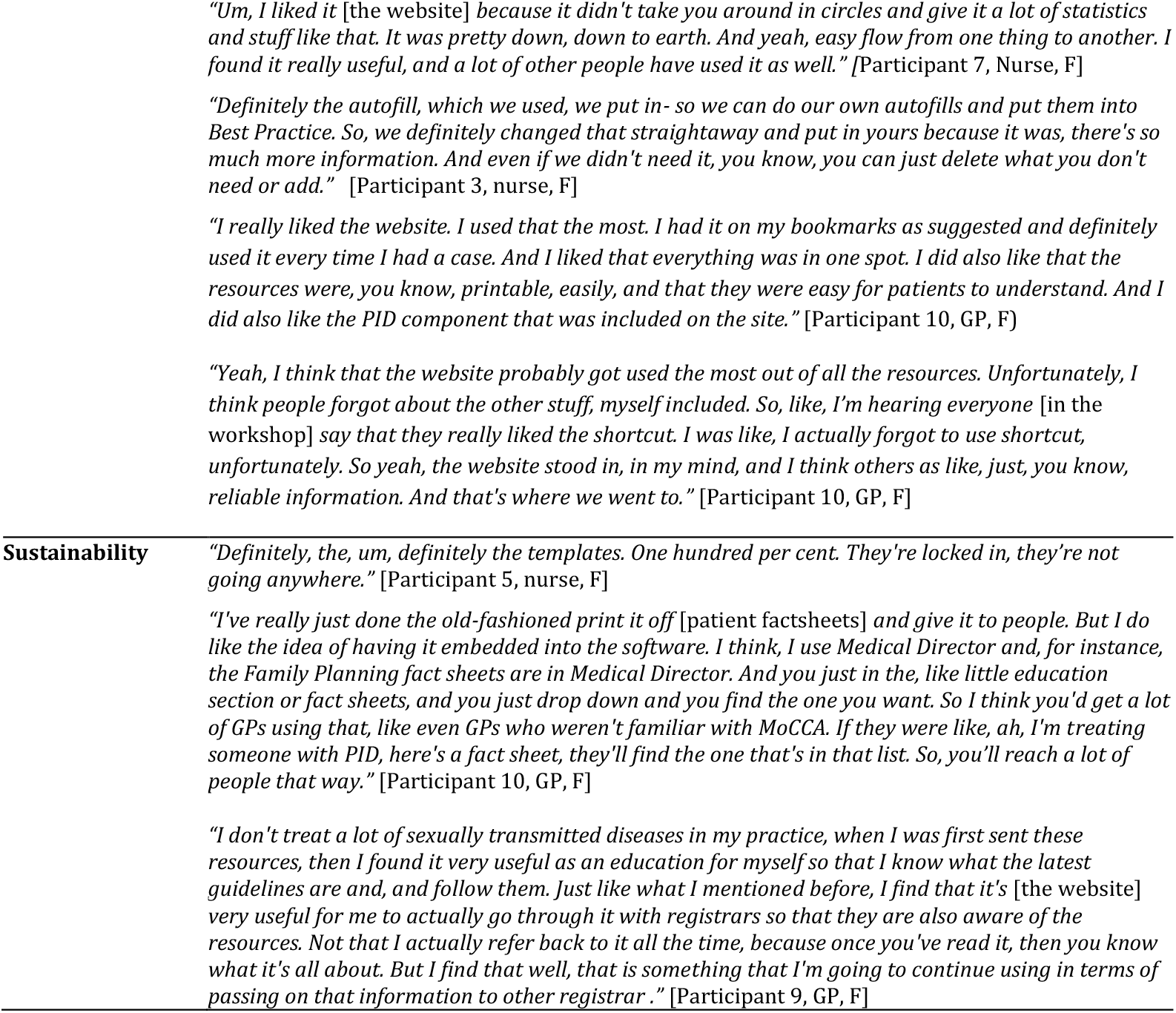
Supporting quotes for MoCCA implementation outcomes.

### Implementation outcomes

Between 9-12 months of the intervention, a total of 33 clinical staff (GPs=26, nurses=7) participated in an interview or workshop (GPs=6, nurses=6), and/or clinic meeting (GPs=22, nurses=5). Our implementation outcome (Box 2) findings are presented below and supporting quotations in table3. Key findings for each MoCCA component are provided in table 4.

**Table 4:** Summary of findings for the MoCCA intervention components.

| Component | How it was used | Positive aspects | Negative aspects |
| --- | --- | --- | --- |
| Website | As an introduction to the MoCCA study<br>In consultation/s for patient/s with a chlamydia infection as a resource to guide patient management, to locate patient resources, and answer patient questions<br>For clinician education about chlamydia management | Relevant information in one place<br>Step-by-step guide that followed STI management guidelines<br>Clear and simple layout<br>Easy to navigate<br>Reliable information<br>Links to useful external resources (e.g. State / Territory information)<br>Time saving | Difficult to find with a Google search<br>Colour scheme not liked by all |
| Shortcuts | To document clinical notes for a chlamydia or PID consultation<br>Not used by some clinicians due to forgetting, lack of opportunity | Clear and comprehensive information.<br>Supports consistency in care and ensures that everything is covered<br>Improves efficiency (time saver)<br>Easy to use and edit | Ease of navigation could be enhanced by including the ^ symbol<br>Does not auto populate medication prescribed |
| Patient factsheets | Printed hard copy or sent electronically to patients<br>Placed on clinic website<br>Not used due to having other factsheets as a default or lack of opportunity | Simple, clear language<br>Well laid out, one page<br>Saved clinician from writing out information<br>Patients liked them<br>Short summary box gave patient key information | Challenging for some to send electronically to patients |
| PDPT resources | To link to PDPT guidance<br>To support PDPT decision making<br>To provide a PDPT prescription<br>Not used by some clinicians due to forgetting, lack of opportunity | Gave clinicians confidence to provide PDPT<br>Informed clinicians that PDPT was allowable in some jurisdictions<br>Nicely formatted | None reported |
| CPD activities | Not used as clinicians had already obtained CPD points | Good to have as an option | None reported |
| Postal retesting | No uptake | Clinicians and patients liked the idea of postal kits. | Awkward to set up and use.<br>Offered to patients but declined.<br>Pre-existing processes (e.g. e-referrals) preferred. |

Overall MoCCA was viewed as an approach that facilitated general practice to provide best practice chlamydia care and that was relevant to other STIs, particularly gonorrhoea and syphilis. The intervention was largely viewed as **acceptable** to help streamline and support chlamydia management and compatible (**appropriateness**) with the general practice setting given that it fitted with existing work processes. An important attribute was that MoCCA reflected best practice as per Australian STI management guidelines, and simplified chlamydia management through a step-by-step approach. Some components (e.g. website, shortcuts, factsheets, PDPT resources) were preferred for their ease of use and support for decision making over other components (e.g. postal retesting kits) which were described as somewhat awkward to organise.

Intention to use MoCCA (**adoption)** and its actual uptake **(penetration)** varied between clinicians and MoCCA components. For example, clinicians often reported using the website, documentation shortcuts, and patient factsheets but not postal retesting or CPD activities. A few clinicians used the PDPT resources. Some clinicians indicated they intended to use MoCCA when they first learned about it but did not ultimately use it as they lacked opportunity to do so (e.g. no patients with a chlamydia diagnosis), had preferred pre-existing practices for chlamydia management (e.g. e-referrals to support retesting rather than postal kits), or forgot. MoCCA was largely set up as intended (**fidelity)** but was not used uniformly. MoCCA was flexible in design, allowing clinicians to use its components in a manner that best suited their needs (e.g. the website was used in consultations to check correct care and educate patients by some clinicians and used out of consultations for education by others).

Importantly, clinicians reflected that MoCCA facilitated best practice chlamydia management (**feasibility)** and that chlamydia management practices had improved for individual clinicians and more broadly in their clinic during the intervention. This included improved continuity of care particularly between GPs and nurses and improved quality of care including more attention to follow up of chlamydia cases such as organising retesting, supporting partner notification or consideration of PID. They also reported improved communication with patients about chlamydia treatment (e.g. shortcuts/factsheets reminded them to discuss retesting/partners). In general, these improvements were viewed as time efficient, in that MoCCA supported better use of clinician’s time.

Many clinicians indicated that they would like MoCCA components to be available for use beyond the trial **(sustainability)** and this was often irrespective of whether they had routinely used them. The website, shortcuts, factsheets and PDPT resources were of particular interest and viewed as a useful approach for other STIs. Preferred avenues for supporting continuation of MoCCA included organisations with STI-care responsibilities such as specialist services or professional organisations. Others noted they would like to see resources embedded in the medical record software.

### Elements influencing implementation

Our directed content analysis across the **CFIR** showed that the attributes of MoCCA (**intervention characteristics)** strongly influenced its implementation and use. The MoCCA intervention was often viewed as providing a **relative advantage** for chlamydia care over existing practices and an approach to providing best practice (**evidence strength and quality)**. Ease of use (**complexity)** and **adaptability** of many MoCCA components to a clinic/clinician needs were important. Some areas to improve the look and feel (**design quality and packaging)** of MoCCA (e.g. website colours) were noted. At an individual level (**individual characteristics)** clinicians’ belief that MoCCA reflected best practice (**knowledge and beliefs)** and that it supported clinical decision making (**self-efficacy)** supported adoption.

Regarding the clinic environment (**inner setting)** a desire to support best practice and improve patient outcomes were motivators to involvement (**implementation climate)** and a practice champion (GP, nurse or practice manager) helped drive and sustain engagement (**readiness for implementation – leadership engagement)**. Relevant external factors (**outer setting**) included that MoCCA was underpinned by best practice and linked directly to known and useful external organisations and resources (**external policy and incentives)**. Furthermore, some clinicians identified opportunities to enhance accessibility of patient resources such as for patients who did not want hard copy information (**patient needs and resources)**.

For the CFIR **process** domain, practice champions (often a nurse) played a key role in promoting and using MoCCA throughout their clinic (**engaging, executing**). For example, putting MoCCA on the agenda for clinic meetings. Clinicians reflected (**reflecting and evaluating)** on ease of use of MoCCA and its value in supporting improved chlamydia management.

Our directed content analysis across the **NPT** complemented our CFIR findings. Many clinicians understood (**coherence)** that MoCCA gave a mechanism to deliver best practice (**cognitive participation),** for example, a reminder of why retesting is important and provided resources to support discussions with patients. GPs and nurses worked together **(collective action)** to use MoCCA when caring for patients with chlamydia, such as using the PDPT prescription template. Clinicians reflected **(reflexive monitoring)** on the usefulness of MoCCA, with many outlining more consistency in their clinic and a more quality consultation.

### Impact outcomes

The proportion of patients with a positive chlamydia test who were retested within 2-4 months of their positive test was 18.1% (n=42/232) pre-intervention and 23.1% (n=75/324) during the intervention (absolute difference = 5.0%, 95% Confidence Interval (CI) −2.1, 12.1) (table 5). Multivariable Poisson regression models showed a significant interaction between intervention period and patient age for retesting within 2-4 months (p=0.02) and analyses were subsequently stratified by age. Adjusting for gender and clinic rurality, retesting for 16–25-year-olds (n=293) increased between the pre-intervention and intervention period (adjusted Relative Risk (aRR) 2.12, 95% CI 1.43, 3.15) and for individuals aged 26-44 years (n=263) results were inconclusive (aRR 1.01, 95% CI 0.66, 1.55) (supplemental information). The overall aRR without stratification by agegroup was 1.37 (95% CI 0.93, 2.02). The intracluster correlation coefficient (ICC) for clustering by clinic for the outcome of retesting within 2-4 months was estimated to be 0.091 (95% CI: 0.001, 0.227).

**Table 5.**
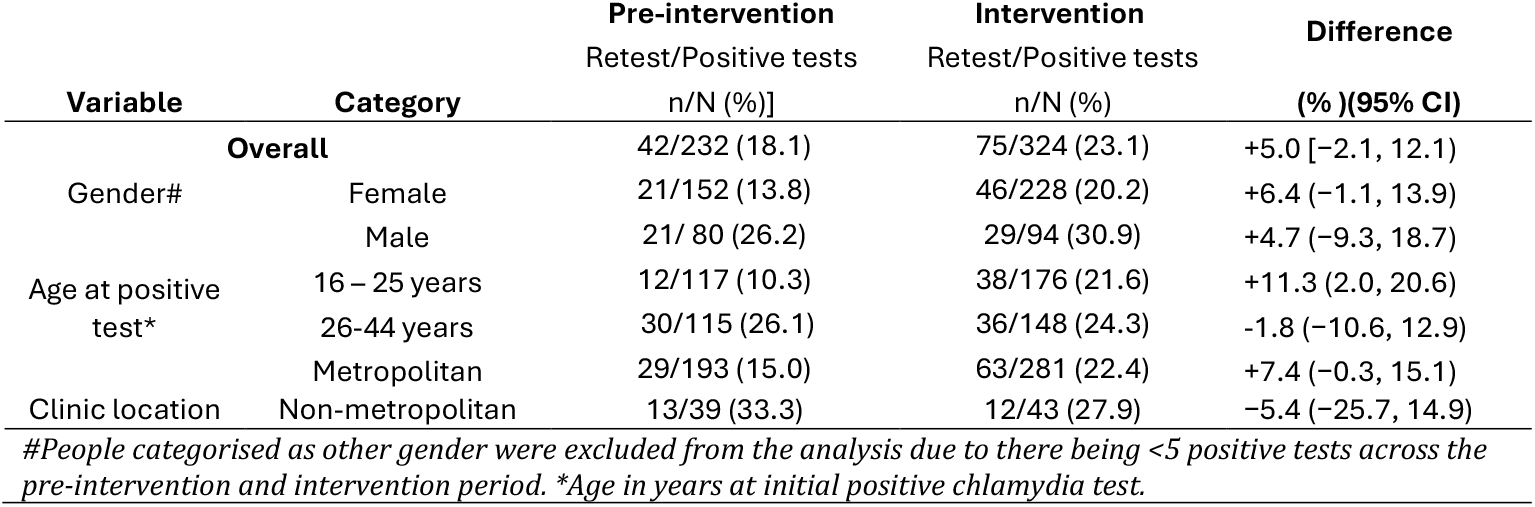
Proportions retested within 2-4 months of a positive test, by intervention period.

Within 2 months of a positive test. 27.2% of all patients were retested in the pre-intervention period and 18.5% of all patients were retested during the intervention period (absolute difference = −8.6%, 95% CI −15.8, 1.5) (supplementary information).

Among patients retested within 2-4 months, 16.7% (95%CI 7.8-31.4) tested positive pre-intervention and 17.3% (95%CI 10.0, 27.7) during the intervention (difference=0.6%, 95%CI 13.8,15.0).

Among female patients, 542 (16.90 per 1000 patients) were diagnosed with PID pre-intervention and 766 (19.08 per 1000 patients) in the intervention period, (absolute difference=2.18 per 1000 patients (95% CI 0.13, 4.23) (table 6). PID rates per 1000 patients were higher for younger patients (16-25 years: pre-intervention= 21.26, 95%CI 18.80, 23.90; intervention= 23.13, 95% CI 20.77,25.68) than for older patients (26+ years: pre-intervention= 14.30, 95% CI 12.72, 16.01; intervention=16.92, 95% CI 15.16, 18.36). Most (95%) PID cases were probable PID. Multivariable Poisson regression models showed a significant interaction between intervention period and clinic location (p=0.02) and were subsequently stratified by clinic location. Adjusting for age, PID diagnoses did not differ between the pre-intervention and intervention period in metropolitan clinics (aRR 1.03, 95% CI 0.69, 1.53) but increased in non-metropolitan clinics (aRR 1.67, 95% CI 1.45, 1.92). The ICC for clustering by clinic for the outcome of PID rates was estimated to be 0.059 (95% CI 0.014, 0.128).

**Table 6:**
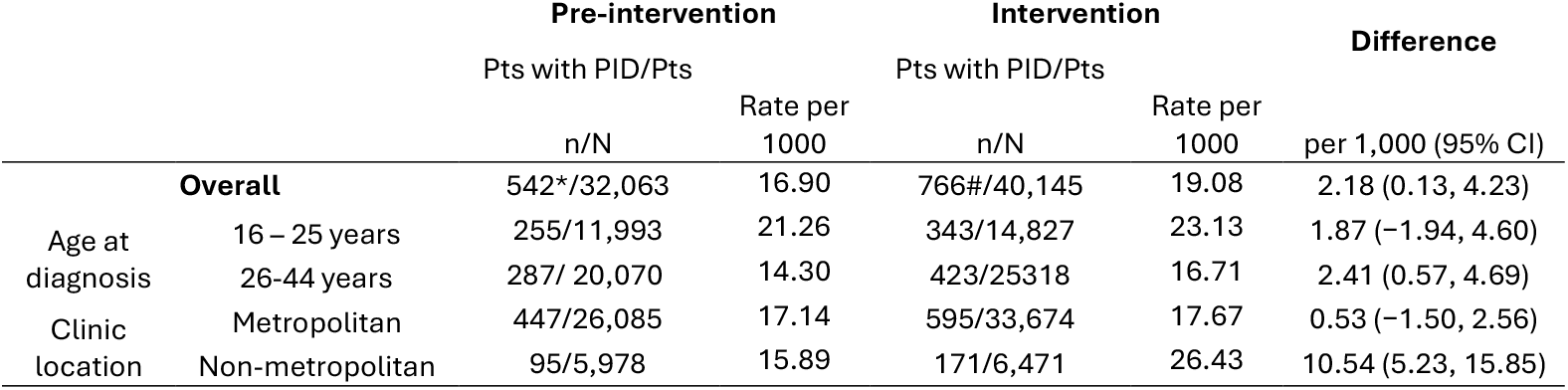
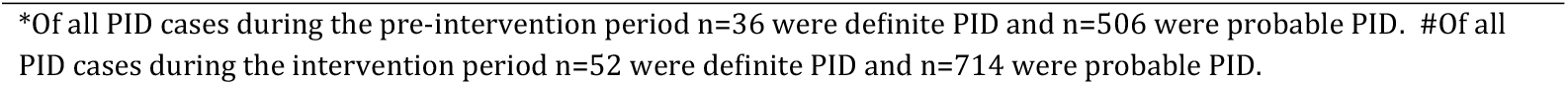
PID rates per 1000 patients, by intervention period.

|  |  | Pre-intervention |  | Intervention |  | Difference<br>per 1,000 (95% CI) |
| --- | --- | --- | --- | --- | --- | --- |
|  |  | Pts with PID/Pts |  | Pts with PID/Pts |  |  |
|  |  | n/N | Rate per<br>1000 | n/N | Rate per<br>1000 |  |
|  | Overall | 542*/32,063 | 16.90 | 766#/40,145 | 19.08 | 2.18 (0.13, 4.23) |
| Age at<br>diagnosis | 16 – 25 years | 255/11,993 | 21.26 | 343/14,827 | 23.13 | 1.87 (–1.94, 4.60) |
|  | 26-44 years | 287/ 20,070 | 14.30 | 423/25318 | 16.71 | 2.41 (0.57, 4.69) |
| Clinic<br>location | Metropolitan | 447/26,085 | 17.14 | 595/33,674 | 17.67 | 0.53 (–1.50, 2.56) |
|  | Non-metropolitan | 95/5,978 | 15.89 | 171/6,471 | 26.43 | 10.54 (5.23, 15.85) |

\*Of all PID cases during the pre-intervention period n=36 were definite PID and n=506 were probable PID. #Of all PID cases during the intervention period n=52 were definite PID and n=714 were probable PID.

## Discussion

### Principal findings

The MoCCA intervention provided general practice clinicians with a clear step-by-step approach and usable resources for best practice chlamydia management. Preferred components were the website, documentation shortcuts, patient information sheets and PDPT resources. Most components aligned well with the general practice setting, were flexible in design, time efficient to use, facilitated decision making, and supported improved chlamydia management including better follow up of chlamydia cases, continuity of care and communication with patients. Quantitative data showed an increase in timely re-testing for younger people, increased PID diagnosis for some groups, and, a decline in retesting too early (<2 months). These are important findings for improving chlamydia case management to reduce the complications of infection. Clinicians generally indicated they would like access to MoCCA resources beyond the study and suggested that organisations with STI-care responsibilities were appropriate avenues.

### Strengths and limitations

A key strength of MoCCA is that it was a partnership project, in which the 11 partner organisations bought extensive expertise and networks in sexual health and general practice research, policy, and care. These attributes ensured the developed MoCCA intervention aligned with general practice and STI control priorities for Australia and that it integrated with the general practice workflow, thereby offering potential for sustainability beyond the project. A further strength is that MoCCA collected comprehensive implementation data that facilitated understandings of how MoCCA was setup, used, and its impact on chlamydia management. The MoCCA approach to implementing an intervention in general practice provides a template that is adaptable to other STIs and conditions and a valuable example of how resources can be integrated into this setting.

MoCCA had several limitations. Firstly, the intervention was at clinic level, but study engagement varied between clinicians and improvements in chlamydia management may not have occurred uniformly. For example, some clinics and clinicians managed few chlamydia cases, thereby limiting opportunity to use MoCCA. However, where used, MoCCA appears to have supported improved quality and continuity of chlamydia management and for clinicians to update their knowledge about chlamydia and STI care. Secondly, we were unable to conduct a randomised trial (RCT) evaluation to assess the effectiveness or cost-effectiveness of MoCCA. Australian general practice experienced significant pressures due to the COVID-19 pandemic, leading to our evaluation of MoCCA in an implementation and feasibility trial. Whilst our sample of 14 clinics was underpowered, our methodology allowed us to collect a rich data set to understand how MoCCA resources were implemented and their quantitative and qualitative impact on chlamydia management. The proportion of patients tested at MoCCA clinics (∼17.5%) was higher than that reported earlier (<10%) for Australian general practice ^6, 24^ whilst positivity (∼5%) was lower than in other reports (∼8%), noting that some earlier reports focussed on individuals aged <30 rather than 16-44 years as was the case in this study. Higher positivity among young males attending general practice than for females has also been reported.^21^ Furthermore, our retesting analysis was limited by the low number of retests. However, although underpowered, our stratified multivariable models did show increased retesting for younger individuals.

### Comparison with other studies

Strengthening capacity for sexual health and STI care in mainstream primary care is a long-standing priority for Australia^10, 41^ and elsewhere.^42^ A range of interventions (many multifaceted) toward increasing such capacity have been evaluated, although it can be difficult to determine the most impactful components of multifaceted interventions. In Australia, a cluster RCT of a chlamydia testing intervention in primary care^6^ showed that audit and feedback were more effective than financial incentives in sustaining increased testing.^43^ A recent rapid review of evidence from multiple countries including Australia, United Kingdom and the USA found that initiatives that target the clinician level (e.g. knowledge and capacity of the general practice workforce such as education, quality improvement, financial incentives) along with clinic-level initiatives (e.g. streamlined work processes) were most effective at increasing STI/HIV testing.^44^

Similarly, MoCCA comprised clinician level components that supported knowledge and clinical decision making (e.g. the website) and clinic level components that supported the practicalities of providing best practice chlamydia management (e.g. documentation shortcuts). In other evidence, models of care that increase the nurses’ role^42^ or link to specialist support^45^ have helped build capacity for STI testing and care in busy general practice environments. At MoCCA clinics, nurses involvement in chlamydia care included support and discussion about partner management, retesting and asking about PID symptoms. The MoCCA shortcuts and other components were viewed as supporting improved continuity of care between GPs and nurses.

### Implications

The three main areas of chlamydia management on which MoCCA focused were partner management, timely retesting for reinfection and PID detection. We found an increase in retesting for people aged 16-25 years, a priority age-group of focus for STI testing.^23^ MoCCA supported improvements in quality and continuity of chlamydia management. GPs and nurses reported a greater focus on partner management, retesting and/or PID symptoms when caring for a patient with chlamydia, as well as the risk of other STIs. Key components emphasised were the website, shortcuts and patient factsheets, with some clinicians favouring PDPT resources. For retesting, clinicians often liked the idea of postal kits, but there was little uptake due to some clinicians viewing them as more awkward to use than their pre-existing retesting processes. In recent years, and during the COVID-19 pandemic, many Australian pathology providers established electronic pathology requests,^46^ that have helped to streamline requests to patients and delivery of results to GPs. This mechanism could be promoted more broadly as part of routine practice for chlamydia and other STIs.

Over recent years, Australia has experienced a concerning increase in syphilis and gonorrhoea diagnoses with women and heterosexual men increasingly affected.^47, 48^ Re-emergence of congenital syphilis and the development of anti-microbial resistant gonorrhoea are urgent concerns with Australian guidelines recommending comprehensive STI testing for chlamydia, gonorrhoea, syphilis and HIV.^23^ It is imperative that general practice can easily locate relevant guidance and have resources that support best practice for sexual history taking, testing and management for all STIs. MoCCA has demonstrated how workflow resources can be developed, quickly implemented and integrated to support STI management and that GPs and nurses will adopt STI focused resources if relevant and aligned with the workflow. Importantly, MoCCA supported improvements in quality and continuity of chlamydia management and was viewed as an approach that could be adapted to other STIs, particularly for gonorrhoea and syphilis. For example, if developed, a shortcut for documenting syphilis consultations might support clinicians to better understand syphilis staging, management and where to access specialist support.

The next step is to realise avenues for transition and scale-up of successful MoCCA components to real world use. Our GPs and nurse participants indicated that mainstream organisations with STI care responsibilities were relevant avenues to facilitate this. MoCCA PDPT resources have been translated to updated health department guidance for PDPT in the state of Victoria where they are available to primary care.^49^ Furthermore, many MoCCA resources are now publicly available to clinicians nationally via the Australian STI Management guidelines.^23^ Discussions are also in progress with specialist services to host MoCCA resources and pathology providers to include a link in STI results to the Australian STI guidelines.

### Further considerations

On a broader perspective the relevance of testing for asymptomatic chlamydia infection is increasingly debated amid concerns about antibiotic overuse and evidence that the risk of harms is lower after asymptomatic than symptomatic infection.^50-52^ Currently for Australia, syphilis is a more urgent concern, being recently declared a communicable disease incident of national significance.^53^ Sexually active Australians are advised to get regular sexual health checks, but evidence shows that young Australians have less knowledge about syphilis than about chlamydia or gonorrhoea.^54^ The role of GPs in taking a sexual history and identifying syphilis infections is crucial and provision of resources for supporting general practice in syphilis management and decision making is imperative. MoCCA has shown how general practice can be supported in chlamydia management. These learnings can be applied to syphilis (and other STIs) on a larger scale and inform an RCT evaluation.

## Conclusions

MoCCA integrated with the general practice setting, supported clinical decision making and improvements in chlamydia management.

## Data Availability

The data that support the findings are not publicly available due to ethical approval for the patient attendance and clinical data pertaining to its collection specifically for the purposes of this study. All relevant data for the present work are contained in the manuscript.

