## Supplemental information for "Strengthening chlamydia management in Australian general practice using interventions that align with the clinic workflow: implementation and feasibility trial findings"

---

### A: Interview and workshop schedule

1. Could you briefly tell me what you know about the MoCCA study?
2. Can you tell me about yours and the clinics engagement with the study over the past 12 months? *(Did engagement change over time, clinic communication channels, barriers to engagement)*
3. Which components of the intervention have you/the clinic used during the study? *(e.g. website, documentation shortcuts, patient factsheets, PDPT or other partner management resources, postal retesting or other retesting resources, professional development activities or articles). (Could you describe any barriers to using the MoCCA components? Any facilitators?)*
4. How useful did you find the MoCCA components that you used? (*How/when did you use them? What did you like / dislike? What impacts did the components have on your practice?)*
5. For any chlamydia positive patients you've been involved with, are there any MoCCA components they have been interested in/engaged with? (*Could anything about the MoCCA intervention be changed to improve the patient experience?)*
6. How or has using the MoCCA intervention impacted your time when caring for a patient with chlamydia? In what way?
7. Have your chlamydia management practices changed since participating in MoCCA? *(Can you tell me how so? What aspect of MoCCA (if any) has had the greatest impact for your clinical practice?*)
8. Would you like to / do you intend to continue using components of MoCCA after the study concludes? *(Which ones and why?)*
9. Where do you think the best place would be for others/yourself to access MoCCA components after the study concludes? *(eg. professional organisations, other)*
10. Would it be beneficial to use the MoCCA approach for other STIs (eg. gonorrhoea, syphilis)? *(What reasons for your answer? Are there other considerations for this?)*
11. Is thing anything else you would like to add about the study?

---

### B: Supplementary tables

*Supp table 1: Age stratified Poisson models for change in retesting within 2-4 months of a positive test*

| | | RR (95% CI) | aRR (95%CI) |
|---|---|---|---|
| **Patients aged 16-25 years at the time of positive test** | | | |
| Gender | Female | Ref. | Ref. |
| | Male | 0.63 (0.27 - 1.45) | 0.63 (0.26 - 1.54) |
| Clinic location | Metropolitan | Ref. | Ref. |
| | Non-metropolitan | 1.28 (0.32 - 5.07) | 1.46 (0.35 - 6.01) |
| Intervention period | Pre-intervention | Ref. | Ref. |
| | Intervention | 2.12 (1.40 - 3.20) | 2.12 (1.43 - 3.15) |
| **Patients aged 26-44 years at the time of positive test** | | | |
| Gender | Female | Ref. | Ref. |
| | Male | 2.22 (1.59 - 3.09) | 2.11 (1.55 - 2.89) |
| Clinic location | Metropolitan | Ref. | Ref. |
| | Non-metropolitan | 1.58 (0.67 - 3.73) | 1.23 (0.63 - 2.42) |
| Intervention period | Pre-intervention | Ref. | Ref. |
| | Intervention | 0.97 (0.62 - 1.52) | 1.01 (0.66 - 1.55) |

*Supp table 2: Proportions retested within 2 months of a positive test, by intervention period.*

| Variable | Category | **Pre-intervention** Retest/Positive tests n/N (%) | **Intervention** Retest/Positive tests n/N (%) | **Difference** (%)(95% CI) |
|---|---|---|---|---|
| **Overall** | | 63/232 (27.2) | 60/324 (18.5) | −8.6 (−15.8, −1.5) |
| Gender# | Female | 45/152 (29.6) | 41/228 (18.0) | −11.6 (−20.2, −3.0) |
| | Male | 18/80 (22.5) | 19/94 (20.2) | −2.3 (−14.6, 10.0) |
| Age at positive test* | 16 – 25 years | 28/117 (23.9) | 32/176 (18.2) | −5.7 (−14.8, 3.4) |
| | 26-44 years | 34/115 (29.6) | 27/148 (18.2) | -11.4 (−21.5, −1.3) |
| Clinic location | Metropolitan | 51/193 (26.4) | 53/ 281 (18.9) | −7.5 (−14.5, −0.5) |
| | Non-metropolitan | 12/39 (30.8) | 7 / 43 (16.3) | −14.5 (−30.8, 1.8) |

*#People categorised as other gender were excluded from the retesting analysis due to there being <5 positive tests across the pre-intervention and intervention period. *Age in years at initial positive chlamydia test.*
